# NATURAL HISTORY OF LAFORA DISEASE A Prognostic Systematic Review and Individual Participant Data Meta-Analysis

**DOI:** 10.1101/2021.06.17.21259096

**Authors:** Federica Pondrelli, Lorenzo Muccioli, Laura Licchetta, Barbara Mostacci, Corrado Zenesini, Paolo Tinuper, Luca Vignatelli, Francesca Bisulli

**Author notes:** **Corresponding author**: Francesca Bisulli, IRCCS Institute of Neurological Sciences of Bologna, Italy, Department of Biomedical and Neuromotor Sciences, University of Bologna, Bellaria Hospital, Via Altura 3 Bologna, Italy 40139. These authors contributed equally (Luca Vignatelli, Francesca Bisulli).

## Abstract

**Objective:** To describe the clinical course of Lafora Disease (LD) and identify predictors of outcome by means of a prognostic systematic review with individual participant data meta-analysis.

**Methods:** A search was conducted on MEDLINE and Embase with no restrictions on publication date. Only studies reporting genetically confirmed LD cases were included. Kaplan-Meier estimate was used to assess probability of death and of loss of autonomy. Univariable and multivariable Cox regression models with mixed effects (clustered survival data) were performed to evaluate prognostic factors.

**Results:** Seventy-three papers describing 298 genetically confirmed LD cases were selected. Mean age at disease onset was 13.4 years (SD 3.7), with 9.1% aged≥ 18 years. Overall survival rates in 272 cases were 93% [95% CI 89-96] at 5 years, 62% [95% CI 54-69] at 10 years and 57% [95% CI 49-65] at 15 years. Median survival time was 11 years. The probability of loss of autonomy in 110 cases was 45% [95% CI 36-55] at 5 years, 75% [95% CI 66-84] at 10 years, and 83% [95% CI 74-90] at 15 years. Median loss of autonomy time was 6 years. Asian origin and age at onset <18 years emerged as negative prognostic factors, while type of mutated gene and symptoms at onset were not related to survival or disability.

**Conclusions:** This study documented that half of patients survived at least 11 years. The notion of actual survival rate and prognostic factors is crucial to design studies on the effectiveness of upcoming new disease-modifying therapies.

## INTRODUCTION

Lafora disease (LD) is a rare and severe autosomal recessive progressive myoclonus epilepsy^1^ (OMIM#254780).

First described in 1911 by Gonzalo Rodriguez-Lafora, LD has a worldwide prevalence close to four cases per million^2^. It is more frequent in Mediterranean countries, North Africa, the Middle East and, overall, in countries with high consanguinity rates^3^.

At present, more than one hundred causative mutations involving two genes, *EPM2A* (6q24) and *EPM2B/NHLRC1* (6p22.3), have been identified as responsible for more than 90% of LD cases^1^. A third gene, *PRDM8* (4q21.21), has been tentatively linked to a new form of early-onset LD^4^. To date, however, no follow-up studies have confirmed its role.

*EPM2A* and *EPM2B* gene products, laforin and malin respectively, form an enzymatic complex involved in several neuronal metabolic pathways, including glycogen metabolism, heat shock response and protein degradation^5^. Laforin or malin loss of function results in polyglucan accumulation in different tissues, such as brain, muscle, liver and skin. Targeted genetic testing is currently the reference standard to confirm the diagnosis, whereas skin biopsy, which might reveal the pathognomonic Lafora bodies, is fraught with false positive and false negative results^5^. The clinical manifestations of LD are primarily due to pathologic neuronal storage of polyglucan. The disease course is characterized by disabling myoclonus, intractable seizures and dementia, as well as ataxia and visual manifestations, resulting in complete loss of autonomy at later stages of the disease. Death is traditionally thought to occur within ten years of onset, mainly related to status epilepticus, aspiration pneumonia or other complications common in chronic neurodegenerative diseases^1,6,7,8,9^.

Possibly due to the rarity of the disease, the natural history of LD and its prognostic factors have not yet been systematically investigated. As in other rare diseases, it is almost impossible to perform single-centre cohort studies thus, in the absence of data from international registries, one option is the aggregation of data from case reports/case series^10^. In this setting, individual participant data meta-analysis^11,12,13^ may be an appropriate methodological approach for summarizing data, also from a prognostic perspective.

Even though no specific treatment for LD is available, promising new therapeutic strategies are currently being tested in animal models and will hopefully soon be available for clinical trials^14^. Therefore, there is a crucial need to establish reference parameters for use in evaluating the real impact, on disease duration and quality of life, of upcoming treatments for LD.

We thus performed a systematic prognostic review with individual participant data meta-analysis of all genetically confirmed LD cases reported in the literature, aiming to better define the disease course and possibly identify prognostic factors.

## METHODS

### Search Strategy

This study was conducted in compliance with the reporting guidelines for prognostic systematic reviews^15^ and individual participant data meta-analysis^10^. A protocol was registered in the PROSPERO database (CRD42020190877). A systematic literature search of the PubMed/MEDLINE and Embase databases was performed, using various combinations of specific key terms (Table 1).

**Table 1.**
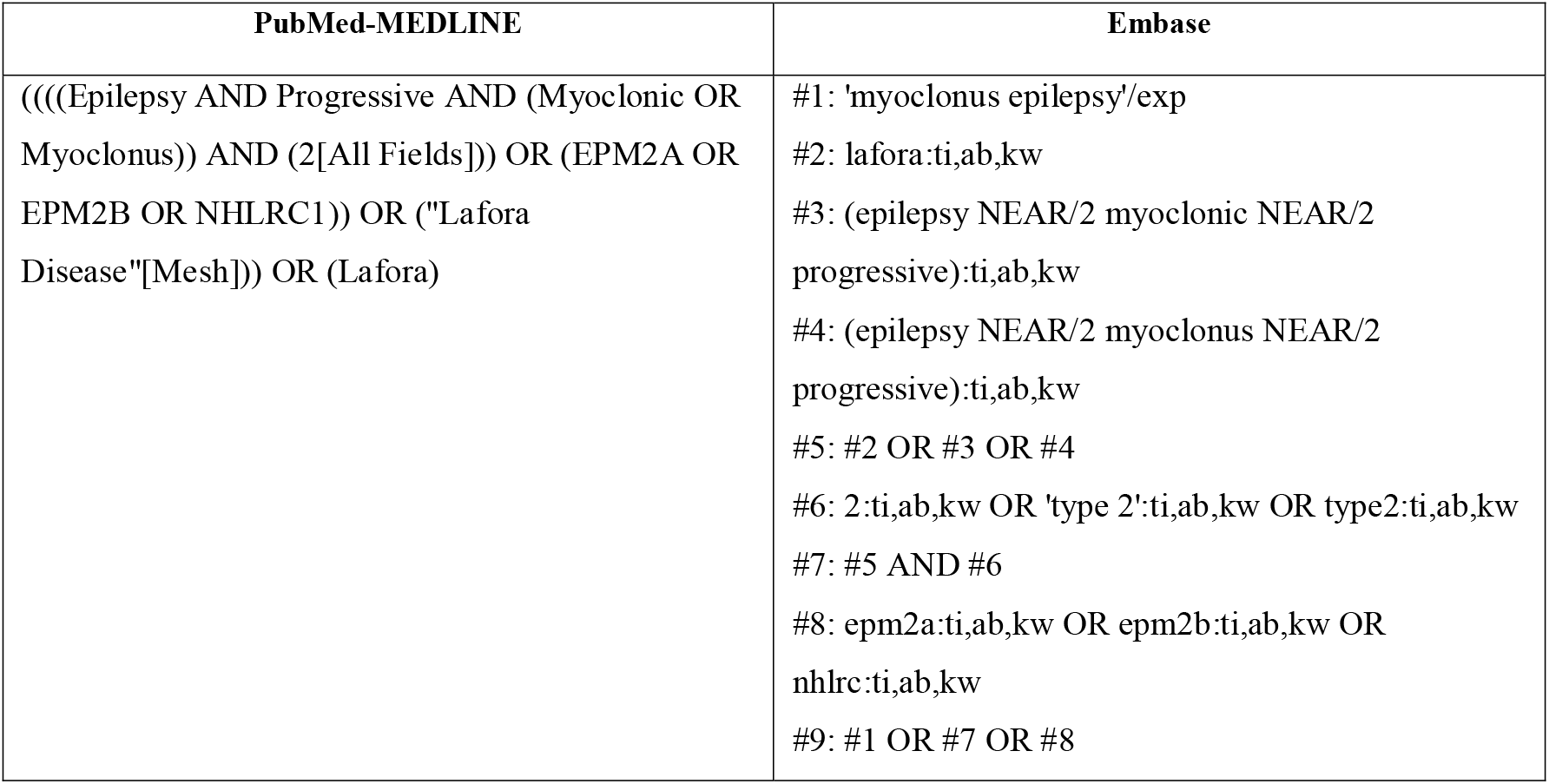
Search Strategy.

| PubMed-MEDLINE | Embase |
| --- | --- |
| (((Epilepsy AND Progressive AND (Myoclonic OR Myoclonus)) AND (2[All Fields])) OR (EPM2A OR EPM2B OR NHLRC1)) OR ("Lafora Disease"[Mesh])) OR (Lafora) | #1: 'myoclonus epilepsy'/exp<br>#2: lafora:ti,ab,kw<br>#3: (epilepsy NEAR/2 myoclonic NEAR/2 progressive):ti,ab,kw<br>#4: (epilepsy NEAR/2 myoclonus NEAR/2 progressive):ti,ab,kw<br>#5: #2 OR #3 OR #4<br>#6: 2:ti,ab,kw OR 'type 2':ti,ab,kw OR type2:ti,ab,kw<br>#7: #5 AND #6<br>#8: epm2a:ti,ab,kw OR epm2b:ti,ab,kw OR nhlrc:ti,ab,kw<br>#9: #1 OR #7 OR #8 |

There was no restriction on the publication date. The last search was performed in June 2021. One reviewer (FP) selected relevant papers through title, abstract and full-text screening. The reference lists of the identified articles were also reviewed to find additional references.

### Eligibility criteria

Eligible study designs included original reports of individual or aggregate data regarding LD patients, published in the form of case reports and case series, while reviews and concept papers were excluded. In cases of overlapping data, the most recent and comprehensive study was considered. Only patients with genetically confirmed LD, *i*.*e*. those harbouring biallelic pathogenic mutations in *EPM2A* or *EPM2B*, were included. We excluded cases diagnosed solely based on skin biopsy considering its poor sensitivity and specificity^6^, or clinical features, and cases with negative genetic test results. In addition to genetic confirmation, a description of the disease history (at least age at onset) or data on disease duration at last follow up were required for inclusion.

Finally, we excluded cases harbouring pathogenic mutations of both *EPM2A* and *EPM2B*, because these rare cases could not be included in a specific genetic category^16,17^.

### Data extraction and management

An *ad hoc* database was created to collect the following information: author, publication year, study type, demographic data, geographical origin of the family/case (if this was not explicitly stated, the country was assumed to be that of the first author’s institutional affiliation), presence of consanguinity, LD clinical history (age at first neurological manifestation, age at onset of the main clinical features namely seizures, myoclonus, visual manifestations and mental deterioration, age at loss of autonomy, age at death or last observation), EEG findings and genetic testing results. Two independent reviewers (FP, LM) evaluated the selected reports and extracted the data mentioned above concerning every single case described. Any disagreements concerning the interpretation of patient data were resolved by discussion and, if necessary, by seeking the opinion of a third reviewer (FB).

Three main categories of clinical presentation were distinguished based on the most significant features at disease onset:

- onset with epilepsy, if seizures (excluding myoclonic ones) alone were reported;
- motor onset, if myoclonus or cerebellar signs, alone or in combination with seizures, were reported (we considered myoclonus a feature separate from seizures, since its pathogenesis in progressive myoclonic epilepsies is still unclear)^18^;
- cognitive onset, if cognitive disturbances (in terms of school difficulties or behavioural changes), alone or in combination with seizures and/or motor symptoms, were reported.

To systematically evaluate disability progression in LD, we examined the disease course descriptions focusing on psychomotor deterioration to identify the age at loss of autonomy. We considered loss of autonomy as equivalent to grade 3 of the disability scale developed by Franceschetti et al.^19^ This scale is based on the residual motor and mental functions, daily living and social abilities. Grade 3 consists of severe mental and motor impairment, *i*.*e*. need for help in walking, regular assistance in daily living activities and poor social interaction.

In cases of missing or aggregated data, we contacted the corresponding authors directly to obtain the required information.

### Quality Assessment of Individual Studies

Given the lack of tools for evaluating the bias risk of case reports and case series, we used items from the Newcastle-Ottawa scale that were appropriate for our systematic review^20^. From this scale, we removed items relating to comparability and adjustment (because our selected studies were non-comparative) and retained items that focused on case selection, case representativeness and ascertainment of outcome. We were thus left with four items which took the form of the following binary-response questions:

- Did the patient(s) represent the medical centre’s entire case load? (Answer on the basis of the medical centre’s scientific impact on LD).
- Was the diagnosis correctly made? (Answer based on genetic testing).
- Was the follow-up long enough for the outcomes to occur? (Consider death as the principal outcome and an adequate follow-up duration as one in which at least half of the cases reached that outcome).
- Is/Are the case(s) described in detail? (Consider the description to be detailed if at least age at onset AND type of onset were reported).

The quality of a report was considered good when all four criteria were met, moderate when three were met, and poor when two or less were met. The same two reviewers assessed the quality of all the included studies and any disagreements were resolved by discussion.

### Statistical analysis

For the descriptive analysis, continuous variables were presented as mean ± standard deviation (SD), and categorical variables as absolute frequency and relative frequency (%).

The Kaplan-Meier estimate was used to calculate the cumulative time-dependent probability of death or loss of autonomy. The time of entry into the analysis was taken as the year of onset, while the time of the endpoint was the year of death or of loss of autonomy, or the year of the last follow-up information (truncated at 15 years of follow up), whichever came first.

Univariable and multivariable Cox regression models with mixed effects (clustered survival data) were performed in order to study the association between disease duration or time to loss of autonomy and prognostic factors. The analysis was performed using data at single patient level. The included studies were considered in the models as cluster variables^21^.

The following parameters were evaluated as possible predictors of survival and/or loss of autonomy: geographical origin, sex, presence of consanguinity, age at onset (defined as “typical” if <18 years; “late” if ≥18 years), type of onset (defined as “onset with epilepsy”, “motor onset” or “cognitive onset”, as described above), mutated gene (*EPM2A*; *EPM2B*) and compound heterozygosity. The results are presented as hazard ratios (HR) and 95% confidence intervals (95% CI). The assumption of proportional hazard was assessed by Schoenfeld residuals (p> 0.05). Statistical analysis was performed with the Stata SE statistical package, version 14.2.

## RESULTS

The process of identification, screening and selection of eligible articles is described in the form of a PRISMA flow diagram (Figure 1).

**Figure 1.**
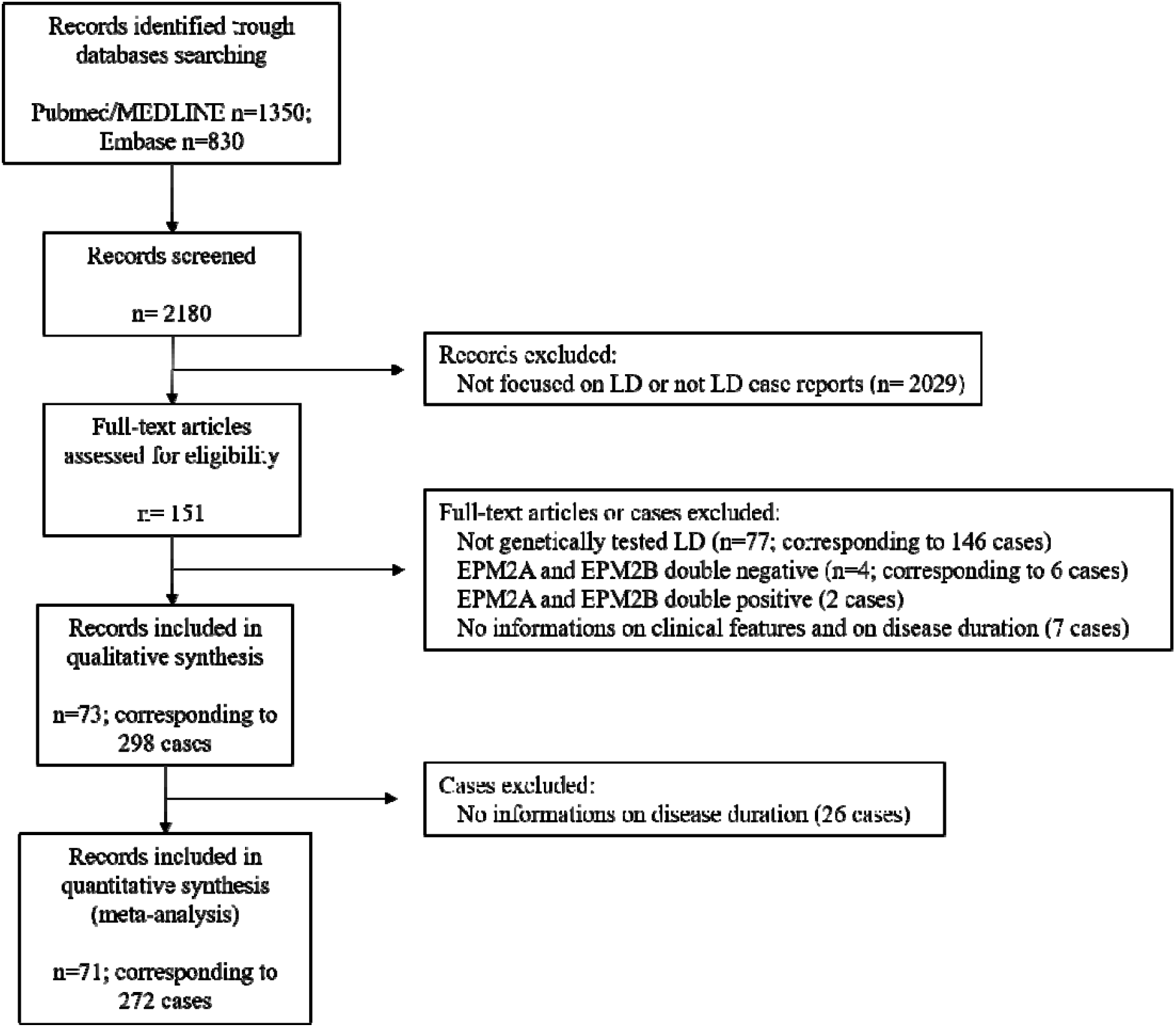
PRISMA Flow Diagram.

Overall, 73 publications corresponding to 298 genetically confirmed cases were eligible for inclusion in the final analysis. Of the 73 papers, 45 described single cases and 28 included two or more cases. The corresponding authors of the 11 studies not reporting complete data on survival were contacted^16,22,23,24,25,26,27,28,29,30,31^: 10 replied that the requested data were unavailable^22,23,24,25,26,27,28,29,30,31^, while one author^16^ provided requested information on one patient.

Thus, 272 cases (91.3%) for which data on disease duration were available were included in the analysis of survival and prognostic factors, while the remaining 26 (8.7%) were included only in the descriptive analysis. Table 2 summarizes the demographic and clinical features of the included patients.

**Table 2.** Demographic and Clinical Features of LD cases.

| Characteristics | n/N (%) or Mean (SD) [range], yr | Characteristics | n/N (%) or Mean (SD) [range], yr |
| --- | --- | --- | --- |
| **Sex, male** | 89/214 (41.6%) | **Visual symptoms** | 60/298 (20.1%) |
| **Geographic Origin** | | Mean age at symptom onset | 12.9 (±2.3) [8.5-18] in 46 |
| European | 154/298 (51.7%) | Mean time from disease onset | 0.5 (±1.0) [0-4] in 46 |
| Asian | 94/298 (31.5%) | **Cognitive decline** |  |
| American | 44/298 (14.8%) | Absent | 11/257 (4.3%) |
| African | 6/298 (2.0%) | Mean age at symptom onset | 15.3 (±5.4) [4-45] in 176 |
| **Family History** | | Mean time from disease onset | 2.3 (±3.6) [0-26] in 176 |
| Number of families/cases | 248/298 | **Mutated Gene** |  |
| Consanguinity | 90/192 (46.9%) | *EPM2A* | 132/298 (44.3%) |
| **Age at disease onset** |  | Compound heterozygosity | 29/132 (22.0%) |
| Mean | 13.4 (±3.7) [4-30] in 298 | *NHLRC1* | 166/298 (55.7%) |
| <18 years | 271/298 (90.9%) | Compound heterozygosity | 41/166 (24.7%) |
| ≥18 years | 27/298 (9.1%) | **Skin Biopsy** | |
| **Type of disease onset** |  | Performed | 138/298 (46.3%) |
| Seizures alone | 149/248 (60.1%) | Positive | 120/138 (86.9%) |
| Motor | 63/248 (25.4%) | **Loss of autonomy** |  |
| Cognitive | 36/248 (14.5%) | Absent | 33/177 (18.6%) |
| **Myoclonus** | | Mean age at onset | 19.4 (±6.2) [10-42] in 83 |
| Absent | 4/246 (1.6%) | Mean time from disease onset | 6.7 (±5.1) [0.2-23] in 83 |
| Mean age at symptom onset | 14.8 (±3.1) [8-28] in 169 | **Dead at last follow up** | 70/272 (25.7%) |
| Mean time from disease onset | 1.0 (±2.0) [0-17] in 169 | Mean age at death | 21.6 (±6.1) [14-59] in 70 |
| **Cerebellar symptoms** | | Mean disease duration | 8.2 (±5.3) [2-40] in 70 |
| Absent | 19/165 (11.5%) |  |  |
| Mean age at symptom onset | 16.7 (±3.4) [10.5-30] in 77 | | |
| Mean time from disease onset | 4.3 (±3.4) [0-14] in 77 | | |

*n/N= number of cases in which a certain characteristic is present out of the total number of cases which it was described; SD= standard deviation.*

The mean age at disease onset was 13.4 years (SD 3.7) [4-30] in 298 subjects, with 27/298 (9.1%) aged ≥ 18 years at onset. As regards the clinical manifestations at onset, 149/248 cases (60.1%) presented with seizures alone, while 63/248 (25.4%) with myoclonus and/or cerebellar signs (alone or in combination with seizures) and 36/248 (14.5%) with cognitive symptoms (alone or in combination with seizures and/or motor symptoms). Visual symptoms, at any stage of the disease, were reported in 60/298 cases (20.1%). As regards genetics, *EPM2A* was mutated in 132/298 cases (44.3%) and *EPM2B* in 166/298 (55.7%). The mean age at loss of autonomy was 19.4 years (SD 6.2) [10-42] in 83 cases. Considering the deceased patients, 70/272 (25.7%), the mean age at death was 21.6 (SD 6.1) [14-59] and the mean disease duration was 8.2 (SD 5.3) [2-40].

### Quality Assessment of Included Studies

Of the 73 publications included in our analysis, 22 (30.2%) were rated as low quality (2 points), 43 (58.9%) as moderate quality (3 points), and 8 (10.9%) as high quality (4 points).

### Survival and Prognostic Factors

Overall survival rates were 93% [95% CI 89-96] at 5 years, 62% [95% CI 54-69] at 10 years and 57% [95% CI 49-65] at 15 years. Considering the lower limit of the 95% CI of the survival curve, the median survival time was 11 years (see Figure 2). Univariable analysis (Table 3) revealed that late-onset (≥18 years) was related to a longer survival [HR 0.44; 95% CI 0.23-0.85]. Multivariable analysis (Table 3) corroborated ≥ 18 years of age at onset as a positive prognostic factor. Asian and America origin emerged as associated to a shorter survival.

**Table 3.** Factors Associated with Shorter Survival.

| Phenotypic Characteristic<br>Variable vs Reference Category | Unadjusted HR<br>(95% CI) | P-value | Adjusted HR (95% CI) | P-value |
| --- | --- | --- | --- | --- |
| **Geographic Origin** |  |  |  |  |
| Asian vs European | 3.5 (1.3-9.2) | **0.011** | 5.0 (1.8-13.4) | **0.001** |
| African vs European | 3.4 (0.4-25.3) | 0.24 | 3.6 (0.5-24.9) | 0.20 |
| American vs European | 2.4 (0.8-7.3) | 0.11 | 3.2 (1.03-9.6) | **0.044** |
| **Sex** |  |  |  |  |
| Male vs Female | 1.4 (0.7-2.6) | 0.27 |  |  |
| **Age at onset** |  |  |  |  |
| ≥ 18 vs < 18 years | 0.44 (0.23-0.85) | **0.014** | 0.21 (0.06-0.79) | **0.021** |
| **Consanguinity** |  |  |  |  |
| Present vs Absent | 1.5 (0.7-3.0) | 0.31 |  |  |
| **Mutated Gene** |  |  |  |  |
| *EPM2B* vs *EPM2A* | 1.1 (0.5-2.5) | 0.75 | 1.3 (0.5-3.0) | 0.59 |
| **Compound Heterozygosity** |  |  |  |  |
| Present vs Absent | 0.8 (0.4-1.7) | 0.63 |  |  |
| **Type of Onset** |  |  |  |  |
| Motor vs Epileptic | 1.9 (0.9-4.1) | 0.091 | 1.4 (0.7-3.0) | 0.36 |
| Cognitive vs Epileptic | 1.0 (0.4-4.0) | 0.96 | 0.8 (0.3-2.0) | 0.59 |

**Figure 2.**
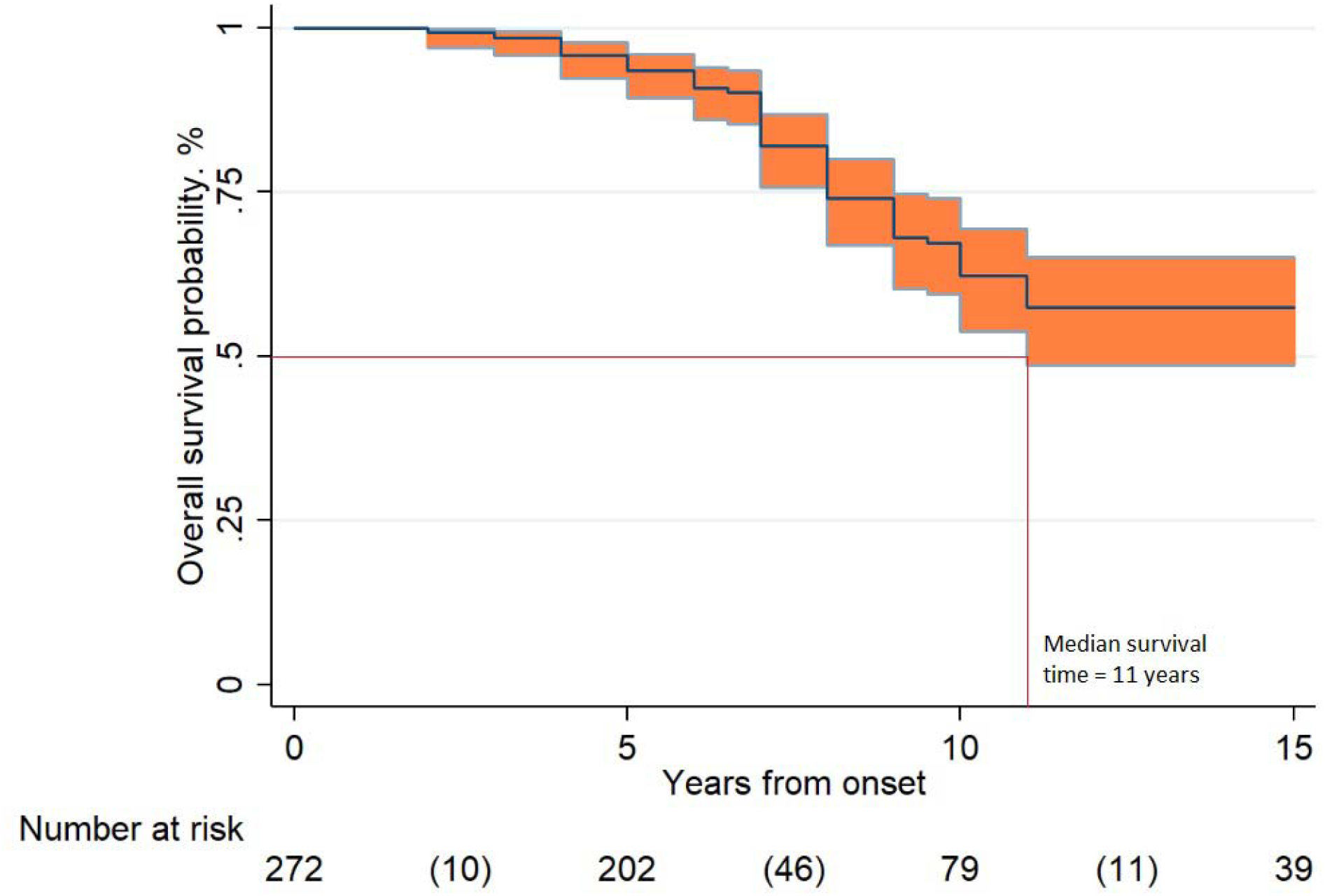
Overall Survival. Legend: Overall survival probability in 272 LD cases according to Kaplan-Meier analysis. The overall survival rates resulted 93% [95% CI 89-96] at 5 years, 62% [95% CI 54-69] at 10 years and 57% [95% CI 49-65] at 15 years (between parentheses the number of events in the time intervals).

**Figure 3.**
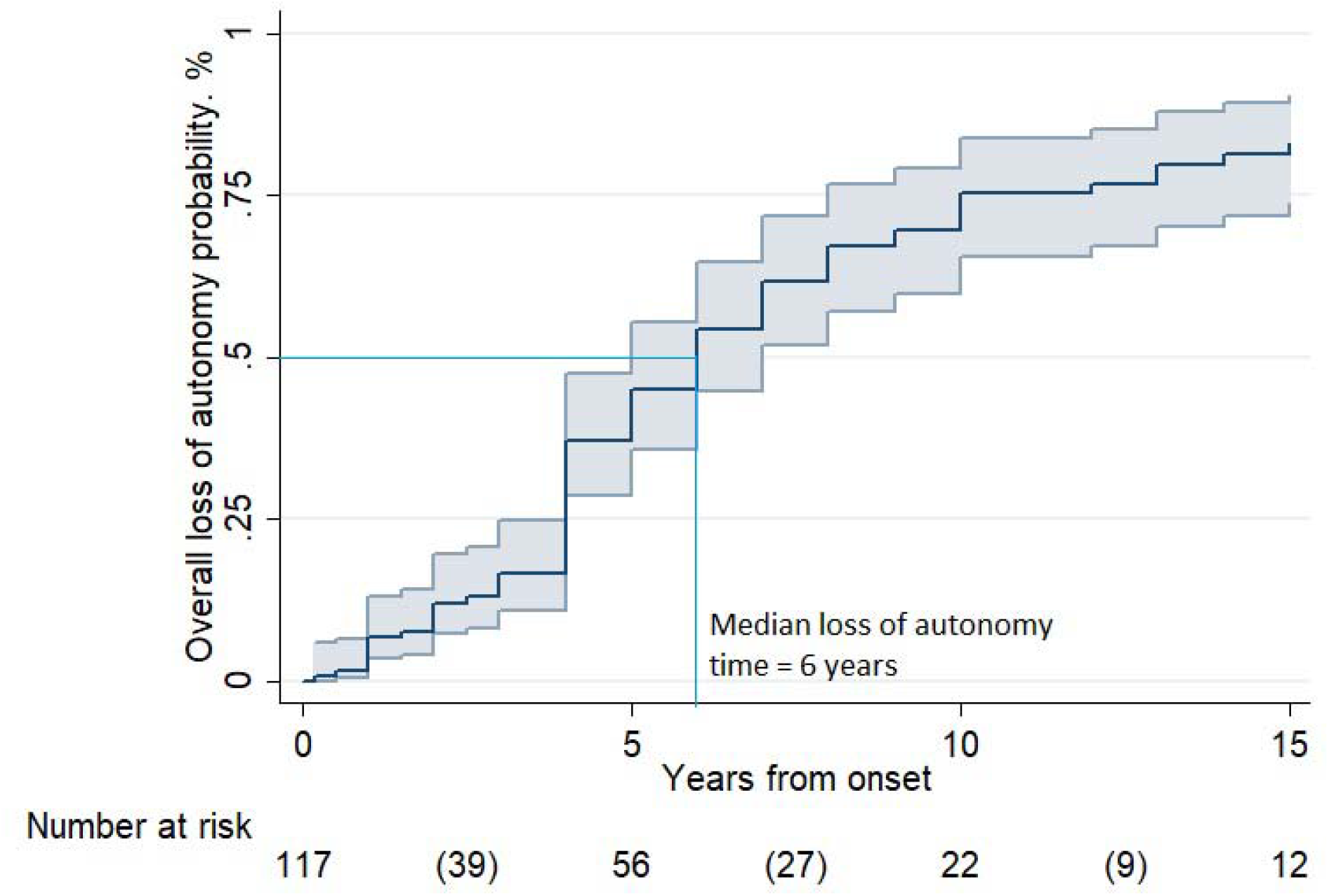
Loss of autonomy. Legend: Overall probability of loss of autonomy in 110 LD cases, according to Kaplan-Meier analysis. The probability of loss of autonomy resulted 45% [95% CI 36-55] at 5 years, 75% [95% CI 66-84] at 10 years, and 83% [95% CI 74-90] at 15 years (between parentheses the number of events in the time intervals).

### Loss of Autonomy and Prognostic Factors

The probability of loss of autonomy was 45% [95% CI 36-55] at 5 years, 75% [95% CI 66-84] at 10 years, and 83% [95% CI 74-90] at 15 years. The median loss of autonomy time was 6 years in the whole group and in the group of patients with age onset <18 years. In those with late-onset (≥18 years) it was 8 years. Multivariable analysis (Table 4) revealed that disability progression differed significantly according to geographical origin and age at onset: Asian patients showed a shorter time to loss of autonomy [HR 4.0; 95% CI 1.3-12.1], while late-onset (≥18 years) [HR 0.20; 95% CI 0.04-0.88] was related to a slower psychomotor deterioration.

**Table 4.** Factors Associated with Shorter Time from Disease Onset to Loss of Autonomy.

| Phenotypic Characteristic<br>Variable vs Reference Category | Unadjusted HR<br>(95% CI) | P-value | Adjusted HR (95% CI) | P-value |
| --- | --- | --- | --- | --- |
| **Geographic Origin** |  |  |  |  |
| Asian vs European | 2.5 (0.9-7.0) | 0.071 | 4.1 (1.4-12.7) | **0.013** |
| African vs European | 7.4 (0.3-180) | 0.222 | 7.4 (0.2-284) | 0.28 |
| American vs European | 1.8 (0.7-4.7) | 0.20 | 2.3 (0.8-6.8) | 0.13 |
| **Sex** |  |  |  |  |
| Male vs Female | 1.0 (0.4-2.8) | 0.95 |  |  |
| **Age at onset** |  |  |  |  |
| ≥ 18 vs < 18 years | 0.48 (0.24-0.96) | **0.039** | 0.18 (0.04-0.79) | **0.024** |
| **Consanguinity** |  |  |  |  |
| Present vs Absent | 1.2 (0.5-2.5) | 0.71 |  |  |
| **Mutated Gene** |  |  |  |  |
| *EPM2B* vs *EPM2A* | 2.7 (0.9-8.6) | 0.084 | 1.6 (0.4-7.4) | 0.53 |
| **Compound Heterozygosity** |  |  |  |  |
| Present vs Absent | 0.8 (0.4-1.6) | 0.52 |  |  |
| **Type of Onset** |  |  |  |  |
| Motor vs Epileptic | 0.9 (0.3-3.1) | 0.93 | 0.9 (0.3-3.0) | 0.82 |
| Cognitive vs Epileptic | 0.5 (0.2-1.6) | 0.26 | 0.7 (0.2-2.0) | 0.49 |

## DISCUSSION

The present systematic study describes the natural history of LD and investigates the prognostic value of demographic and clinical features in a large sample of genetically confirmed cases published in the literature.

Analysis unexpectedly showed that at least 50% of patients survived 11 years (median survival time), suggesting that the disease course could be longer than previously reported.

The notion that affected individuals usually die within ten years of onset is often reported in the existing literature^1,6,7,8,9^. This statement derived from the earliest studies, mainly based on autoptic diagnosis^32^, before genetic testing became available. Moreover, the available reports are narrative, non-systematic reviews, in which it is possible that only the most severe and/or peculiar cases were selected. Thus, our finding may be explained considering several factors such as applying a systematic approach that allowed collection of a larger and more representative sample; the advent of molecular diagnosis enabling early detection of even the milder cases, and the improvement of supportive care.

Investigation of disability progression revealed that 50% of the patients lost autonomy within 6 years of onset (median loss of autonomy time). This is a potentially important observation to design the upcoming new drugs evaluation protocols correctly.

Indeed, the aim of a disease-modifying therapy in LD should be twofold, on the one hand prolonging survival, but also delaying disability progression. Another important consequence of this finding is that, as subjects with a rapid disability progression are largely represented among patients with LD, their exclusion from therapeutic trials aimed at merely prolonging survival would significantly narrow the eligible population.

Concerning prognostic factors, late-onset (≥18 years) appeared to be related to longer disease duration and also to slower progression to loss of autonomy. It could be speculated that more prolonged survival is due to slower accumulation of Lafora bodies (LBs) and/or to a more favourable distribution of LBs in the central nervous system.

Geographical origin also emerged as a prognostic factor, with patients from Asia found to have a poorer prognosis, possibly related to genetic factors and, on the other hand, to socioeconomic issues. Of note, studies on epilepsy epidemiology in Asia reported a higher standardized mortality ratio (SMR) in epileptic Asian patients compared to Western populations^33^.

Conversely, symptoms at onset and the type of mutated gene did not seem to correlate with LD prognosis. Regarding genetics, our finding failed to support some reports suggesting that involvement of *EPM2B* generally may be related to slower disease progression^28,29^. However, we propose that phenotypic variations are mainly attributable to specific mutation types and/or to interactions with other “modifier genes”^8^. This is in line with the severe and rapidly progressive phenotypes associated with specific *EPM2B* mutations^34,35,36^ and, on the other hand, slowly progressive forms also associated with specific *EPM2A* mutations^23,37,38^.

Analysis of the population’s overall characteristics revealed that geographical distribution is in line with descriptions of a higher LD prevalence in Mediterranean countries, the Middle East and India^39^.

Our sample showed a wide range of ages at disease onset (4-30 years), suggesting that LD should also be considered in the differential diagnosis of young children and adults presenting with epilepsy and myoclonus.

The clinical manifestations of LD, both at onset and during the disease course, seemed to vary widely from case to case, even though seizures were the first symptom in the majority of patients (60.1%). Visual symptoms are traditionally considered a characteristic feature of LD although the epileptic origin has been debated^40^. In our series, these were reported in only about 20% of the cases. It is plausible that visual manifestations, even if present, were not always mentioned by the authors of the selected studies, as they may not have constituted a crucial element for the purposes of their reports.

### Limitations

Our study is based on single case reports and case series, which are ranked as the lowest level of evidence. However, we applied several methodological tools to explore or minimise the possible sources of bias. For example, we sought to minimise the clustering effect by applying a regression model with mixed effects for clustered survival data. It is also possible that our findings are affected by availability bias, since the reported clinical information differed widely between studies, resulting in fewer observations for some parameters. Moreover, we may have failed to identify some duplicate cases due to the anonymisation of patient data.

The estimates on the duration of survival and the time to loss of autonomy could be inflated by the not negligible number of censored patients in the Kaplan-Meier analysis. However, even in a worst-case scenario (*i*.*e*., assuming that all the patients reported as lost to follow up were actually deceased), at least 20% of patients survive at 10 years and 14% at 15 years (data not shown). Our results may also be affected by publication bias, given the possibility that single case reports with unusual clinical characteristics are the ones more likely to get published. Against that, several of the included studies reported quite sizeable case series.

## Conclusions

This review systematically investigates the natural history of LD in a large sample of genetically confirmed cases. Half of the patients lost autonomy within six years of onset and survived at least eleven years of onset. In addition, we identified age at onset and the patient’s geographical origin as possible prognostic factors. Notably, the type of mutated gene didn’t emerge as a prognostic factor. This study provides preliminary data useful to the design multicentre clinical trials assessing the effectiveness of upcoming disease-modifying therapies.

## Supporting information

Supplemental References

PRISMA checklist

## Data Availability

The authors confirm that the data supporting the findings of this study are available within the article and its supplementary materials.

## Acknowledgments

The authors thank A.I.LA. (Associazione Italiana Lafora; http://www.lafora.it/) for inspiring this work by encouraging a deeper understanding of the subject.

Also thank to Maria Camerlingo, Agenzia sanitaria e sociale regionale, Regione Emilia-Romagna, for assisting with the search strategy and to Prof. Damir Janigro, MD, PhD, Department of Physiology, Case Western Reserve University, Cleveland, OH, USA, for assisting in manuscript translation.

## Competing interests

Authors declare no competing interests.

## Notes

### Competing Interest Statement

The authors have declared no competing interest.

### Funding Statement

This research received no funding.

### Author Declarations

IRB and/or ethics committee approvals were not necessary, since we extracted data from the literature.

