## Supplemental References for "NATURAL HISTORY OF LAFORA DISEASE A Prognostic Systematic Review and Individual Participant Data Meta-Analysis"

**Supplement- References of the included papers**

Afrantou T, Lagoudaki R, Papadopoulos T et al. Novel frameshift variant of NHLRC1 gene in compound heterozygosity in an adult Greek patient with Lafora disease. *Seizure.* 2021 Mar;86:49-51. doi: 10.1016/j.seizure.2021.01.011. Epub 2021 Jan 23.

Ahmad A, Dad R, Ullah MI et al. Clinical and genetic studies in patients with Lafora disease from Pakistan. *J Neurol Sci.* 2017 Feb 15;373:263-267. doi: 10.1016/j.jns.2017.01.010.

Al Mufargi Y, Qureshi A, Al Asmi A. Lafora Disease: Report of a Rare Entity. *Cureus.* 2020 Jan 28;12(1):e6793. doi: 10.7759/cureus.6793.

Altindag E, Kara B, Baykan B et al. MR spectroscopy findings in Lafora disease. *J Neuroimaging.* 2009 Oct;19(4):359-65. doi: 10.1111/j.1552-6569.2008.00325.x.

Andrade DM, del Campo JM, Moro E et al. Nonepileptic visual hallucinations in Lafora disease. Neurology. 2005 Apr 12;64(7):1311-2. doi: 10.1212/01.WNL.0000156907.49247.05.

Andrade, Ackerley CA, Minett TSC et al. Skin biopsy in Lafora disease: genotype-phenotype correlations and diagnostic pitfalls. *Neurology.* 2003 Dec 9;61(11):1611-4. doi: 10.1212/01.wnl.0000096017.19978.cb.

Araya N, Takahashi Y, Shimono M et al. A recurrent homozygous NHLRC1 variant in siblings with Lafora disease. *Hum Genome Var.* 2018 Jul 12;5:16. doi: 10.1038/s41439-018-0015-9.

Arslan EA, Öncel I, Ceylan AC et al. Genetic and phenotypic features of patients with childhood ataxias diagnosed by next-generation sequencing gene panel. *Brain Dev.* 2020 Jan;42(1):6-18. doi: 10.1016/j.braindev.2019.08.004.

Aslam Z, Lee E, Badshah M et al. Whole exome sequencing identified a novel missense mutation in EPM2A underlying Lafora disease in a Pakistani family. *Seizure*. 2017 Oct;51:200-203. doi: 10.1016/j.seizure.2017.08.012.

Baykan B, Striano P, Gianotti S et al. Late-onset and slow-progressing Lafora disease in four siblings with EPM2B mutation. *Epilepsia.* 2005 Oct;46(10):1695-7. doi: 10.1111/j.1528-1167.2005.00272.x.

Begic E, Bradaric H, Begic Z et al. Lafora Disease during a Seven-Year Period, Bosnian and Herzegovinian experience. *Iran J Child Neurol.* Winter 2019;13(1):115-120.

Béjot Y, Lemesle-Martin M, Contégal F et al. Lafora's disease presenting with progressive myoclonus epilepsy. *Revue Neurologique* 2007; 163(10):975-978. doi: https://doi.org/10.1016/S0035-3787(07)92642-9

Bisulli F, Muccioli L, d'Orsi G et al. Treatment with metformin in twelve patients with Lafora disease. *Orphanet J Rare Dis.* 2019 Jun 21;14(1):149. doi: 10.1186/s13023-019-1132-3.

Brackmann FA, Kiefer A, Agaimy A et al. Rapidly progressive phenotype of Lafora disease associated with a novel NHLRC1 mutation. *Pediatr Neurol.* 2011 Jun;44(6):475-7. doi: 10.1016/j.pediatrneurol.2011.01.012.

Brenner D, Baumgartner T, von Spiczak S et al. Genotypes and phenotypes of patients with Lafora disease living in Germany. Neurol Res Pract. 2019;1:34. doi: 10.1186/s42466-019-0040-2.

Çalışkan D, Dündar NO, Karahan N et al. Lafora disease and occipital lobe seizures: case report. *Turkiye Klinikleri J Pediatr* 2014;23(1):36-39.

Cardinali S, Canafoglia L, Bertoli S et al. A pilot study of a ketogenic diet in patients with Lafora body disease. *Epilepsy Res.* 2006 May;69(2):129-34. doi: 10.1016/j.eplepsyres.2006.01.007.

Casciato S, Gambardella S, Mascia A et al. Severe and rapidly-progressive Lafora disease associated with NHLRC1 mutation: a case report. *Int J Neurosci.* 2017 Dec;127(12):1150-1153. doi: 10.1080/00207454.2017.1337012.

Chan EM, Bulman DE, Paterson AD et al. Genetic mapping of a new Lafora progressive myoclonus epilepsy locus (EPM2B) on 6p22. *J Med Genet.* 2003 Sep;40(9):671-5. doi: 10.1136/jmg.40.9.671.

Chatzistefanidis D, Giaka K, Georgiou I et al. A novel nonsense mutation of the EPM2A gene in northwest Greece causing myoclonic epilepsy. *Seizure.* 2013 May;22(4):315-7. doi: 10.1016/j.seizure.2012.12.014.

Corcia L, Hohensee S, Olivero A et al. Lafora disease with novel autopsy findings: a case report with endocrine involvement and literature review. *Pediatr Neurol.* 2014 Nov;51(5):713-6. doi: 10.1016/j.pediatrneurol.2014.07.034.

Couarch P, Vernia S, Gourfinkel-An I et al. Lafora progressive myoclonus epilepsy: NHLRC1 mutations affect glycogen metabolism. *J Mol Med (Berl).* 2011 Sep;89(9):915-25. doi: 10.1007/s00109-011-0758-y.

Dirani M, Nasreddine W, Abdulla F et al. Seizure control and improvement of neurological dysfunction in Lafora disease with perampanel. *Epilepsy Behav Case Rep.* 2014 Sep 29;2:164-6. doi: 10.1016/j.ebcr.2014.09.003.

D'Souza MS, Amirthraj A. Need for interprofessional collaborative practice: Lafora disease

*International Journal of Nutrition, Pharmacology, Neurological Diseases* 2016;6(3):133-135.

El Tahry R, de Tourtchaninoff M, Vrielynck P et al. Lafora disease: psychiatric manifestations, cognitive decline, and visual hallucinations. *Acta Neurol Belg*. 2015 Sep;115(3):471-4. doi: 10.1007/s13760-014-0399-3.

Ferlazzo E, Canafoglia L, Michelucci R et al. Mild Lafora disease: clinical, neurophysiologic, and genetic findings. *Epilepsia.* 2014 Dec;55(12):e129-33. doi: 10.1111/epi.12806. Epub 2014 Sep 30.

Franceschetti S, Gambardella A, Canafoglia L et al. Clinical and genetic findings in 26 Italian patients with Lafora disease. *Epilepsia.* 2006 Mar;47(3):640-3. doi: 10.1111/j.1528-1167.2006.00479.x.

Frantz T, Fortson E, Strowd LC. Utility of Skin Biopsy in a Case of Progressive Myoclonic Epilepsy: Challenge. *Am J Dermatopathol.* 2018 Sep;40(9):e123. doi: 10.1097/DAD.0000000000000924.

Fu Y, Zhou C, Song R et al. A novel compound heterozygous EPM2A mutation in a Chinese boy with Lafora disease. *Neurol Sci.* 2020 Aug;41(8):2267-2270. doi: 10.1007/s10072-020-04377-7.

Ganesh S, Delgado-Escueta AV, Suzuki T et al. Genotype-phenotype correlations for EPM2A mutations in Lafora's progressive myoclonus epilepsy: exon 1 mutations associate with an early-onset cognitive deficit subphenotype. *Hum Mol Genet*. 2002 May 15;11(11):1263-71. doi: 10.1093/hmg/11.11.1263.

Garcia-Gimeno MA, Rodilla-Ramirez PN, Viana R et al. A novel EPM2A mutation yields a slow progression form of Lafora disease. *Epilepsy Res*. 2018 Sep;145:169-177. doi: 10.1016/j.eplepsyres.2018.07.003.

Gökdemir S, Cağlayan H, Kızıltan M et al. Presentation of an unusual patient with Lafora disease. *Epileptic Disord*. 2012 Mar;14(1):94-8. doi: 10.1684/epd.2012.0489.

Goldsmith D, Minassian BA. Extraneurological sparing in long-lived typical Lafora disease. *Epilepsia Open*. 2018 May 17;3(2):295-298. doi: 10.1002/epi4.12224.

Goldsmith D, Minassian BA. Efficacy and tolerability of perampanel in ten patients with Lafora disease. *Epilepsy Behav.* 2016 Sep;62:132-5. doi: 10.1016/j.yebeh.2016.06.041.

Gomez-Abad C, Afawi Z, Korczyn AD et al. Founder effect with variable age at onset in Arab families with Lafora disease and EPM2A mutation. *Epilepsia.* 2007 May;48(5):1011-4. doi: 10.1111/j.1528-1167.2007.01004.x.

Gómez-Abad C, Gómez-Garre P, Gutiérrez-Delicado E et al. Lafora disease due to EPM2B mutations: a clinical and genetic study. *Neurology.* 2005 Mar 22;64(6):982-6. doi: 10.1212/01.WNL.0000154519.10805.F7.

Gómez-Garre P, Gutiérrez-Delicado E, Gómez-Abad C et al. Hepatic disease as the first manifestation of progressive myoclonus epilepsy of Lafora. *Neurology.* 2007 Apr 24;68(17):1369-73. doi: 10.1212/01.wnl.0000260061.37559.67.

González-De la Rosa MG, Alva-Moncayo E. Lafora disease presentation, two cases in a Mexican family. *Rev Med Inst Mex Seguro Soc.* Mar-Apr 2017;55(2):252-256.

Guerrero R, Vernia S, Sanz R et al. A PTG variant contributes to a milder phenotype in Lafora disease. *PLoS One.* 2011;6(6):e21294. doi: 10.1371/journal.pone.0021294.

Hajnsek S, Gadze ZP, Borovecki F et al. Vagus nerve stimulation in Lafora body disease. *Epilepsy Behav Case Rep.* 2013 Sep 27;1:150-2. doi: 10.1016/j.ebcr.2013.08.002.

Harirchian MH, Shandiz EE, Turnbull J et al. Lafora disease: a case report, pathologic and genetic study. *Indian J Pathol Microbiol.* Apr-Jun 2011;54(2):374-5. doi: 10.4103/0377-4929.81645.

Ianzano L, Young EJ, Zhao XC et al. Loss of function of the cytoplasmic isoform of the protein laforin (EPM2A) causes Lafora progressive myoclonus epilepsy. *Hum Mutat.* 2004 Feb;23(2):170-6. doi: 10.1002/humu.10306.

Israni AV, Mandal A. Progressive Myoclonic Epilepsy Due to Lafora Body Disease with a Novel Mutation. *J Pediatr Neurosci.* Jan-Mar 2018;13(1):123-125. doi: 10.4103/JPN.JPN_13_17.

Jara-Prado A, Ochoa A, Alonso ME et al. Late onset Lafora disease and novel EPM2A mutations: breaking paradigms. *Epilepsy Res.* 2014 Nov;108(9):1501-10. doi: 10.1016/j.eplepsyres.2014.08.017.

Kecmanović M, Jović N, Keckarević-Marković M et al. Clinical and genetic data on Lafora disease patients of Serbian/Montenegrin origin. Clin Genet. 2016 Jan;89(1):104-8. doi: 10.1111/cge.12570.

Khiari HM, Lesca G, Malafosse A et al. A novel exon 3 mutation in a Tunisian patient with Lafora's disease. *J Neurol Sci.* 2011 May 15;304(1-2):136-7. doi: 10.1016/j.jns.2011.02.011.

Ki CS, Kong SY, Seo DW et al. Two novel mutations in the EPM2A gene in a Korean patient with Lafora's progressive myoclonus epilepsy. *J Hum Genet*. 2003;48(1):51-4. doi: 10.1007/s100380300006.

Lanoiselée HM, Genton P, Lesca G et al. Are c.436G>A mutations less severe forms of Lafora disease? A case report. *Epilepsy Behav Case Rep.* 2014 Jan 19;2:19-21. doi: 10.1016/j.ebcr.2013.11.003.

Lesca G, Boutry-Kryza N, de Toffol B et al. Novel mutations in EPM2A and NHLRC1 widen the spectrum of Lafora disease. *Epilepsia.* 2010 Sep;51(9):1691-8. doi: 10.1111/j.1528-1167.2010.02692.x.

Lynch DS, Wood NW, Houlden H. Late-onset Lafora disease with prominent parkinsonism due to a rare mutation in EPM2A. *Neurol Genet.* 2016 Aug 16;2(5):e101. doi: 10.1212/NXG.0000000000000101.

Martin S, Strzelczyk A, Lindlar S. Drug-Resistant Juvenile Myoclonic Epilepsy: Misdiagnosis of Progressive Myoclonus Epilepsy. *Front Neurol.* 2019 Sep 10;10:946. doi:10.3389/fneur.2019.00946.

Martínez-Bermejo A, López-Martín V, Serratosa JM et al. Lafora disease. A new case of confirmation of diagnosis on molecular genetic studies. *Rev Neurol.* 2002 Jan 16-31;34(2):117-20.

Mikati MA, Tabbara F. Managing Lafora body disease with vagal nerve stimulation. *Epileptic Disord.* 2017 Mar 1;19(1):82-86. doi: 10.1684/epd.2017.0892.

Mostacci B, Bisulli F, Muccioli L et al. Super refractory status epilepticus in Lafora disease interrupted by vagus nerve stimulation: A case report. *Brain Stimul*. Nov-Dec 2019;12(6):1605-1607. doi: 10.1016/j.brs.2019.08.008.

Nicolescu RC, Al-Khawaga S, Minassian BA et al. Diabetes Mellitus in a Patient With Lafora Disease: Possible Links With Pancreatic β-Cell Dysfunction and Insulin Resistance. *Front Pediatr.* 2019 Jan 16;6:424. doi: 10.3389/fped.2018.00424.

Potes T, Galicchio S, Rosso B et al. Progressive myoclonic epilepsy secondary to Lafora's body disease. *Medicina (B Aires).* 2018;78(6):436-439.

Poyrazoğlu HG, Karaca E, Per H et al. Three patients with lafora disease: different clinical presentations and a novel mutation. *J Child Neurol.* 2015 May;30(6):777-81. doi: 10.1177/0883073814535489.

Ragona F, Canafoglia L, Castellotti B et al. Early Parkinsonism in a Senegalese girl with Lafora disease. *Epileptic Disord.* 2020 Apr 1;22(2):233-236. doi: 10.1684/epd.2020.1150.

Riva A, Orsini A, Scala M et al. Italian cohort of Lafora disease: Clinical features, disease evolution, and genotype-phenotype correlations. *J Neurol Sci.* 2021 May 15;424:117409. doi: 10.1016/j.jns.2021.117409. Epub 2021 Mar 20.

Rudenskaia GE, Zakharova EI, Karpin SL et al. Myoclonic epilepsy of Lafora: a case report. *Zh Nevrol Psikhiatr Im S S Korsakova.* 2010;110(3 Suppl 2):11-6.

Salar S, Yeni N, Gündüz A et al. Four novel and two recurrent NHLRC1 (EPM2B) and EPM2A gene mutations leading to Lafora disease in six Turkish families. *Epilepsy Res.* 2012 Feb;98(2-3):273-6. doi: 10.1016/j.eplepsyres.2011.09.020.

Satishchandra P, Sinha S. Progressive myoclonic epilepsy. *Neurol India*. Jul-Aug 2010;58(4):514-22. doi: 10.4103/0028-3886.68660.

Schorlemmer K, Bauer S, Belke M et al. Sustained seizure remission on perampanel in progressive myoclonic epilepsy (Lafora disease). *Epilepsy Behav Case Rep.* 2013 Aug 16;1:118-21. doi: 10.1016/j.ebcr.2013.07.003.

Singh S, Sethi I, Francheschetti S et al. Novel NHLRC1 mutations and genotype-phenotype correlations in patients with Lafora's progressive myoclonic epilepsy. *J Med Genet.* 2006 Sep;43(9):e48. doi: 10.1136/jmg.2005.039479.

Singh S, Suzuki T, Uchiyama A et al. Mutations in the NHLRC1 gene are the common cause for Lafora disease in the Japanese population. *J Hum Genet.* 2005;50(7):347-352. doi: 10.1007/s10038-005-0263-7.

Striano P, Ackerley CA, Cervasio M et al. 22-year-old girl with status epilepticus and progressive neurological symptoms. Brain Pathol. 2009 Oct;19(4):727-30. doi: 10.1111/j.1750-3639.2009.00308.x.

Striano P, Zara F, Turnbull J et al. Typical progression of myoclonic epilepsy of the Lafora type: a case report. *Nat Clin Pract Neurol.* 2008 Feb;4(2):106-11. doi: 10.1038/ncpneuro0706.

Tee SK, Ong TL, Aris A et al. Lafora disease in a Malaysian with a rare mutation in the EPM2A gene. *Seizure.* 2019 Apr;67:78-81. doi: 10.1016/j.seizure.2019.03.012.

Traoré M, Landouré G, Motley W et al. Novel mutation in the NHLRC1 gene in a Malian family with a severe phenotype of Lafora disease. *Neurogenetics.* 2009 Oct;10(4):319-23. doi: 10.1007/s10048-009-0190-4.

Turnbull J, Kumar S, Ren ZP et al. Lafora progressive myoclonus epilepsy: disease course homogeneity in a genetic isolate. *J Child Neurol.* 2008 Feb;23(2):240-2. doi: 10.1177/0883073807309245.

Vincent A, Macrì A, Tumber A et al. Ocular phenotype and electroretinogram abnormalities in Lafora disease: A "window to the brain". *Neurology.* 2018 Jul 17;91(3):137-139. doi: 10.1212/WNL.0000000000005821.

Yildiz EP, Yesil G, Ozkan MU et al. A novel EPM2A mutation in a patient with Lafora disease presenting with early parkinsonism symptoms in childhood. *Seizure.* 2017 Oct;51:77-79. doi: 10.1016/j.seizure.2017.07.011. Epub 2017 Jul 27.

Zutt R, Drost G, Vos YJ et al. Unusual Course of Lafora Disease. Epilepsia Open. 2016 Aug 25;1(3-4):136-139. doi: 10.1002/epi4.12009.
